# A genome-wide association study identifies genetic variants associated with hip pain in the UK Biobank cohort (N=221,127)

**DOI:** 10.1101/2023.09.20.23295811

**Authors:** Qi Pan, Tengda Cai, Abi Veluchamy, Harry L Hebert, Peixi Zhu, Mainul Haque, Tania Dottorini, Lesley A Colvin, Blair H Smith, Weihua Meng

**Affiliations:** Nottingham Ningbo China Beacons of Excellence Research and Innovation Institute, University of Nottingham Ningbo China, Ningbo, China, 315100; Division of Population Health and Genomics, Ninewells Hospital and Medical School, University of Dundee, Dundee, UK, DD2 4BF; College of Pharmaceutical Sciences, Zhejiang University of Technology, Hangzhou, Zhejiang, China, 310014; School of Mathematical Sciences, University of Nottingham Ningbo China, Ningbo, China, 315100; School of Veterinary Medicine and Science, University of Nottingham, Nottingham, UK, LE12 5RD

**Keywords:** Hip pain, UK Biobank, genetic correlations, genome-wide association study

## Abstract

Hip pain is a common musculoskeletal complaint that leads many people to seek medical attention. Approximately 14.3% of the population aged 60 and above have reported substantial hip pain persisting for most days over a six-week period. Our research aimed to identify the genetic variants associated with hip pain by conducting a genome-wide association study (GWAS) on the hip pain phenotype, utilizing data from 221,127 participants from the UK Biobank cohort. We found 7 different loci associated with hip pain, with the most significant SNP being rs77641763, which is situated within the *EXD3* gene (*p* value = 2.20 x 10^-13^). We utilized publicly available summary statistics from a previous GWAS meta-analysis on hip osteoarthritis as a replication cohort. Two loci (rs12042579 and rs9597759) were suggestively replicated. Further analysis of tissue expression revealed significant associations between brain tissues and hip pain. Additionally, we found strong genetic correlations between hip pain and other pain phenotypes. This research has therefore identified multiple genetic loci associated with hip pain, which may potentially pave the way for medical interventions that can alleviate the burden of hip pain.

## Introduction

Traditionally defined, hip pain denotes any sensation of unease or suffering encountered within the expanse between the upper thigh and the pelvis, frequently emanating from the hip joint itself or the encompassing anatomical structures (Mills et al. 2022). Hip pain is an incapacitating affliction affecting a substantial portion of the global population (Elbrhamy et al. 2023). It can arise from a multitude of underlying causes, spanning from traumatic injuries to degenerative ailments like osteoarthritis (OA), or inflammatory disorders such as bursitis (Kim and Carrier 2022; Pianka et al. 2021).

Understanding the prevalence and determinants of hip pain is vital for healthcare policy-making and targeted interventions. Within a cohort of the US population, 14.3% of individuals aged 60 and above indicated experiencing significant hip pain for most days in a six-week period (Christmas et al. 2002). A higher prevalence was observed among females than among males (Christmas et al. 2002). Given that hip pain is a salient symptom of hip osteoarthritis, the prevalence and characteristics of hip osteoarthritis may, to a certain degree, mirror the manifestations of hip pain. The Framingham Osteoarthritis Study serves as another pivotal cohort study shedding light on hip osteoarthritis prevalence. Initiated in Framingham, Massachusetts in the 1940s, this longitudinal research has played a crucial role in elucidating diverse musculoskeletal disorders, including hip osteoarthritis (Felson 1990). Recent findings from this study suggest that hip osteoarthritis prevalence ranges between 15% and 20% among participants aged 50 and above, with a higher prevalence noted among females and the elderly (Zhang and Jordan 2010). Additionally, the European Project on Osteoarthritis, encompassing population-based cohort studies across 5 European nations, documented a hip osteoarthritis prevalence of approximately 20% among individuals aged between 65 and 74 years (Schaap et al. 2011). To some extent, the wide-ranging disparities in prevalence across countries highlight the importance of socioeconomic and genetic determinants, as well as the need for a consensus on definition in hip pain epidemiology (Schaap et al. 2011).

Numerous epidemiological studies have endeavored to elucidate the risk determinants associated with hip disorders, particularly hip osteoarthritis. High initial body mass index (BMI), sedentary lifestyles, obesity, prior joint injuries, and specific occupational activities have been recognized as factors correlated with hip disorders, mainly hip osteoarthritis (Bierma-Zeinstra and Koes 2007; Blagojevic et al. 2010; Gelber et al. 1999; Lau et al. 2000; Liu et al. 2007; van Dijk et al. 2006; Wang et al. 2022). Beyond environmental determinants, the potential genetic foundations of hip osteoarthritis have garnered significant research attention. Investigations involving familial clusters and twin cohorts have suggested a genetic predisposition to hip osteoarthritis, underscoring the potential importance of genetic components in its initiation and progression (Lindberg 1986; MacGregor and Spector 1999).

While the genetic basis of hip pain has been a topic of interest, genetic studies to date have focused on hip osteoarthritis rather than hip pain more generally. Historically, the genetic architecture of hip osteoarthritis has been perceived to adhere to an additive genetic model, wherein there is an interplay of multiple genes or loci occurs, each exerting a relatively small effect size. Notable contributing genes, such as *COL27A1, MTRF1LP2, CCDC26, DELEC1, TNC, LMX1B* and *SMO*, have been reported as having strong associations with hip osteoarthritis (Boer et al. 2021; Casalone et al. 2018; Styrkarsdottir et al. 2018; Tachmazidou et al. 2019; Zengini et al. 2018; Zhang et al. 2023). Furthermore, a recent GWAS conducted by Henkel et al, including over 700,000 participants, has identified several novel genetic variants that contribute specifically to both non-surgical and surgical hip osteoarthritis (Henkel et al. 2023).

To identify the genetic variants associated with hip pain, we undertook a GWAS employing the expansive UK Biobank cohort. Replication of findings was tested by comparison with the Henkel cohort noted above (Henkel et al. 2023)

## Materials and Methods

### Cohorts’ information

The UK Biobank cohort recruited more than 500,000 individuals aged between 40 and 69 years across England, Scotland, and Wales (2006-2011). The participants completed a detailed questionnaire and standard clinical examination, including health and sociodemographic information, and provided biological samples from which DNA was extracted. They gave informed consent for their health records to be utilized for research purposes. Full details of the UK Biobank cohort are available at their website www.ukbiobank.ac.uk. Ethical approval for the study was given by the UK’s National Health Service National Research Ethics Service (reference 11/NW/0382).

The process of DNA extraction and quality control (QC) was standardized, with comprehensive methodologies available for reference at https://biobank.ctsu.ox.ac.uk/crystal/ukb/docs/genotyping_sample_workflow.pdf. The Wellcome Trust Centre for Human Genetics at Oxford University had responsibility for the standardized QC procedures relating to genotyping outcomes. A thorough description of these QC steps is accessible at http://biobank.ctsu.ox.ac.uk/crystal/refer.cgi?id=155580. In July 2017, the UK Biobank disseminated genetic data — encompassing both directly genotyped and imputed genotypes — from 501,708 specimens for the benefit of sanctioned researchers. The meticulous QC procedures for imputation have been delineated by Bycroft et al (Bycroft et al. 2018).

For this study, a specific pain-related question, created by the UK Biobank, was employed: ‘in the last month have you experienced any of the following that interfered with your usual activities?’ The options were: (1) Headache; (2) Facial pain; (3) Neck or shoulder pain; (4) Back pain; (5) Stomach or abdominal pain; (6) Hip pain; (7) Knee pain; (8) Pain all over the body; (9) None of the above; and (10) Prefer not to say. More than one option could be selected (UK Biobank Questionnaire field ID: 6159). Cases of hip pain in this research were identified by participants choosing the ’hip pain’ response, irrespective of their selections of other categories. Control subjects were distinguished by their selection of the ’None of the above’ option. Data from individuals not of white British descent were excluded from the analysis to mitigate population stratification and increase study homogeneity.

In the replication phase, we used the publicly available summary statistics from Henkel et al’s GWAS meta-analysis on hip osteoarthritis, described above (Henkel et al. 2023). The main reason for this choice was that there are no perfect hip pain cohorts available. Henkel et al’s study sought to discern potential genetic differences between osteoarthritis patients who had undergone joint replacement and those who had not. The research included both surgical hip osteoarthritis cases (N = 20,221) and non-surgical cases (N = 17,847) sourced from Denmark (the Copenhagen Hospital Biobank pain and degenerative musculoskeletal disease study and the Danish Blood Donor Study), Iceland (genetic programmes at deCODE genetics), and the UK Biobank. Within each of the three datasets, primary cases of knee and hip osteoarthritis were identified and categorized into two groups based on whether they had undergone knee/hip replacement surgery. Control groups consisted of individuals with no documented diagnoses of osteoarthritis or rheumatoid arthritis. Given that this is a meta-analytic GWAS study encompassing three datasets, and each SNP could be influenced by any of the three cohorts, we emphasized results that were not influenced by the UK Biobank to enhance the robustness of our research.

### Statistical analysis

In this study, the Genome-wide Complex Trait Analysis (GCTA, v1.94.1) software (available at https://yanglab.westlake.edu.cn/software/gcta/#Overview) served as the primary tool for conducting GWAS (Yang et al. 2011). Our research used the fastGWA function within GCTA for the GWAS analysis, a mixed linear model association tool. Standard QC procedures were applied, entailing the exclusion of single nucleotide polymorphisms (SNPs) with INFO scores below 0.1, SNPs with minor allele frequencies lower than 0.5%, or SNPs failing the Hardy-Weinberg tests (*p* < 10^-6^). Additionally, SNPs situated on the X and Y chromosomes, as well as mitochondrial SNPs, were eliminated from consideration. The association tests were performed using the fastGWA function with adjustments for age, sex, BMI, and eight population principal components. A χ^2^ test was employed to investigate gender differences between the cases and controls, and the comparison of age and BMI was conducted through an independent t-test, using IBM SPSS 22 (IBM Corporation, New York). A *p* value less than 5 x 10^-8^ was considered indicative of genome-wide association significance. Moreover, GCTA was also utilized to compute the narrow-sense heritability.

### GWAS-associated analysis by FUMA and LDSC

The FUMA web application served as the primary annotation tool throughout hip pain study (Watanabe et al. 2017). Additionally, both a Manhattan plot and a Q–Q plot were generated using this application. To offer regional visualization, Locus Zoom (http://locuszoom.org/) was employed (Pruim et al. 2010). Notably, FUMA facilitated three key types of analyses, gene analysis, gene-set analysis, and tissue expression analysis.

In the gene analysis, the summary statistics of SNPs were aggregated to the level of entire genes. This aggregation enabled the assessment of associations between genes and the phenotype under investigation. Gene-set analysis, specific groups of genes sharing common biological, functional, or other characteristics were collectively tested. This approach provided valuable insights into the involvement of distinct biological pathways or cellular functions in the genetic basis of the observed phenotype. The tissue expression analysis was conducted using data from GTEx (https://www.gtexportal.org/home), which has been integrated into the FUMA platform. For this analysis, the average gene expression per tissue type was utilized as a gene covariate. This approach allowed for the examination of relationships between gene expression in specific tissue types and the genetic associations with hip pain which is the focus of the study.

To identify potential genetic correlations between hip pain and other types of pain disorders, we employed linkage disequilibrium score regression through LDSCv1.0.1 (available at https://github.com/bulik/ldsc) (Bulik-Sullivan et al. 2015). Employing the patterns of linkage disequilibrium – the non-random association of alleles at different loci within a population – LDSC provided valuable insights into the potential genetic overlap between distinct traits. This powerful tool shed light on the underlying genetic relationships between hip pain and other pain-related conditions. The statistical data pertaining to other pain-related phenotypes were primarily derived from prior research on multisite pain, knee pain, headache, facial pain, neck or shoulder pain, abdominal pain, and back pain, utilizing the UK Biobank dataset (Johnston et al. 2019; Meng et al. 2019; Meng et al. 2020a; Meng et al. 2020b).

## Results

### GWAS results

During the initial assessment phase (2006-2010) of the UK Biobank study, 501,708 participants were presented with a pain questionnaire. Of these respondents, 49,926 indicated experiencing ’Hip pain’ (hereafter referred to as ’cases’ in this study), while 177,417 chose the ’None of the above’ option (hereafter referred to as ’controls’ in this study). After exclusion of non-white British participants and samples that did not meet QC criteria, we identified 48,563 cases (comprising 18,270 males and 30,293 females) and 172,564 controls (consisting of 81,693 males and 90,871 females) for the GWAS analysis. The study had 11,165,459 SNPs at its disposal for the GWAS examination.

Table 1 provides a detailed overview of the clinical attributes of both the cases and controls. Within the UK Biobank samples, statistical disparities existed between cases and controls with respect to sex, age, and BMI.

**Table 1.** Clinical characteristics of hip pain cases and controls in the UK Biobank.

| Covariates | Cases | Controls | *p*-value |
| --- | --- | --- | --- |
| Sex (male: female) | 18,270:30,293 | 81,693:90,871 | <0.001 |
| Age (years) | 58.5 (7.53) | 57.0 (7.94) | <0.001 |
| BMI (kg/m<sup>3</sup>) | 28.8 (5.36) | 26.7 (4.30) | <0.001 |

A $\chi^2$ test was used to test the difference of gender frequency between cases and controls and an independent t test was used for other covariates.
Continuous covariates were presented as mean (standard deviation)
BMI represents body mass index.

In our study, we identified 7 distinct SNP clusters exhibiting significant associations with hip pain, attaining genome-wide significance *p* < 5 x 10^-8^, as shown in Figure 1. Within the 7 identified loci, Table 2 enumerates a total of 12 independent and significant SNPs, each being the most statistically significant association within its respective locus. A comprehensive list of all significantly associated SNPs from this GWAS is provided in Supplementary Table 1. The paramount association was observed in the SNP cluster located within the *EXD3* gene on chromosome 9q34.3, bearing a *p* value of 2.20 x 10^-13^ for rs77641763. The second most significantly associated cluster was the *RPRD* gene on chromosome 1, presenting the minimal *p* value of 9.87 x 10^-11^ for rs782595679. The regional plot denoting the most significant locus in *EXD3* is presented in Figure 2 and the regional plots for the other six loci are delineated in Supplementary Figures 2 and 3. Furthermore, the Q–Q plot of the GWAS during the discovery phase is depicted in Supplementary Figure 1. The SNP-based heritability for hip pain was determined to be 0.12, with a standard error of 0.02. The p values of the associations of the 12 independent and significant SNPs from the discovery stage were extracted from Henkel et al’s associations with hip OA. The loci located on chromosomes 1 (*p* = 2.3 x 10^-4^ in surgical hip OA cohort, *p* = 1.10 x 10^-3^ in non-surgical hip OA cohort) and 13 (*p* = 9.80 x 10^-3^ in surgical hip OA cohort) were suggestively replicated during the validation phase. Specifically, the locus on chromosome 1 was replicated in both the surgical and non-surgical hip osteoarthritis cohorts, while the locus on chromosome 13 was only replicated within the non-surgical hip osteoarthritis cohort. (Table 2)

**Fig. 1.**
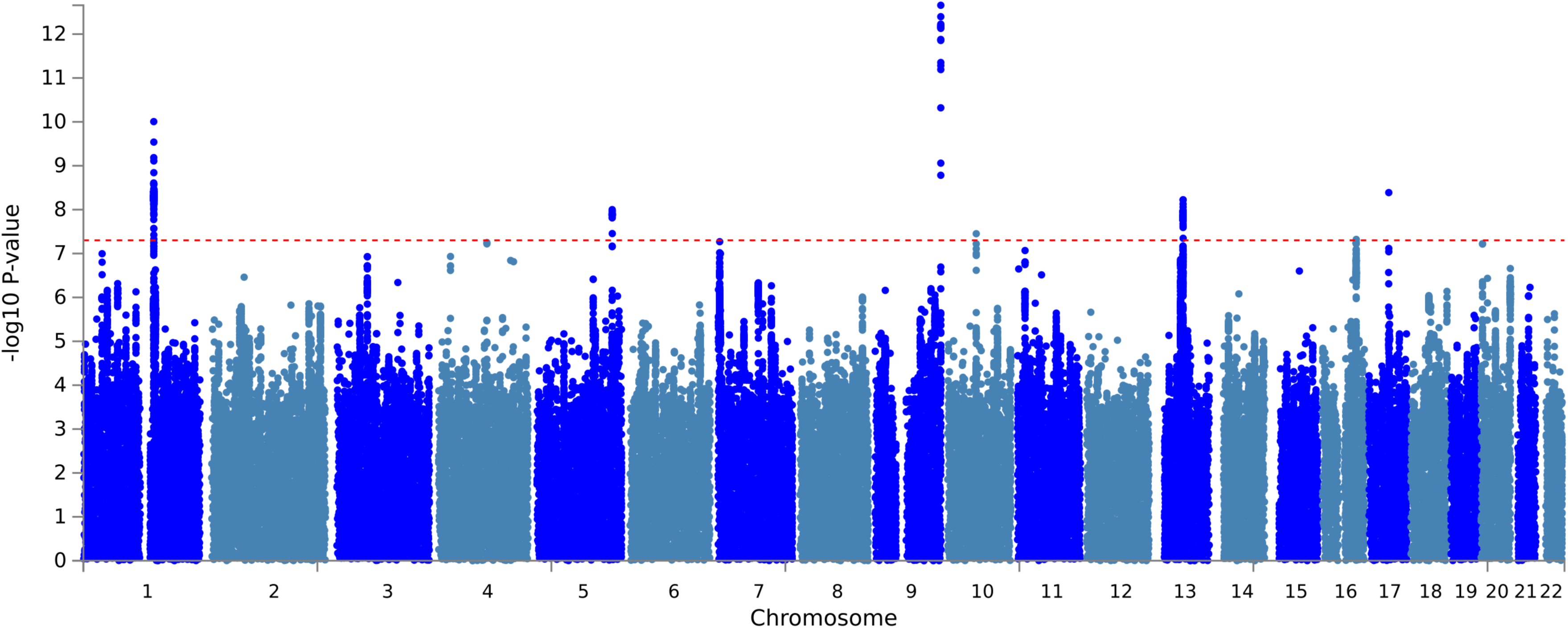
The Manhattan plot of the GWAS analysis on hip pain (N = 221,127) The dashed red line indicates the cut-off *p* value of 5 × 10^−8^

**Fig. 2.**
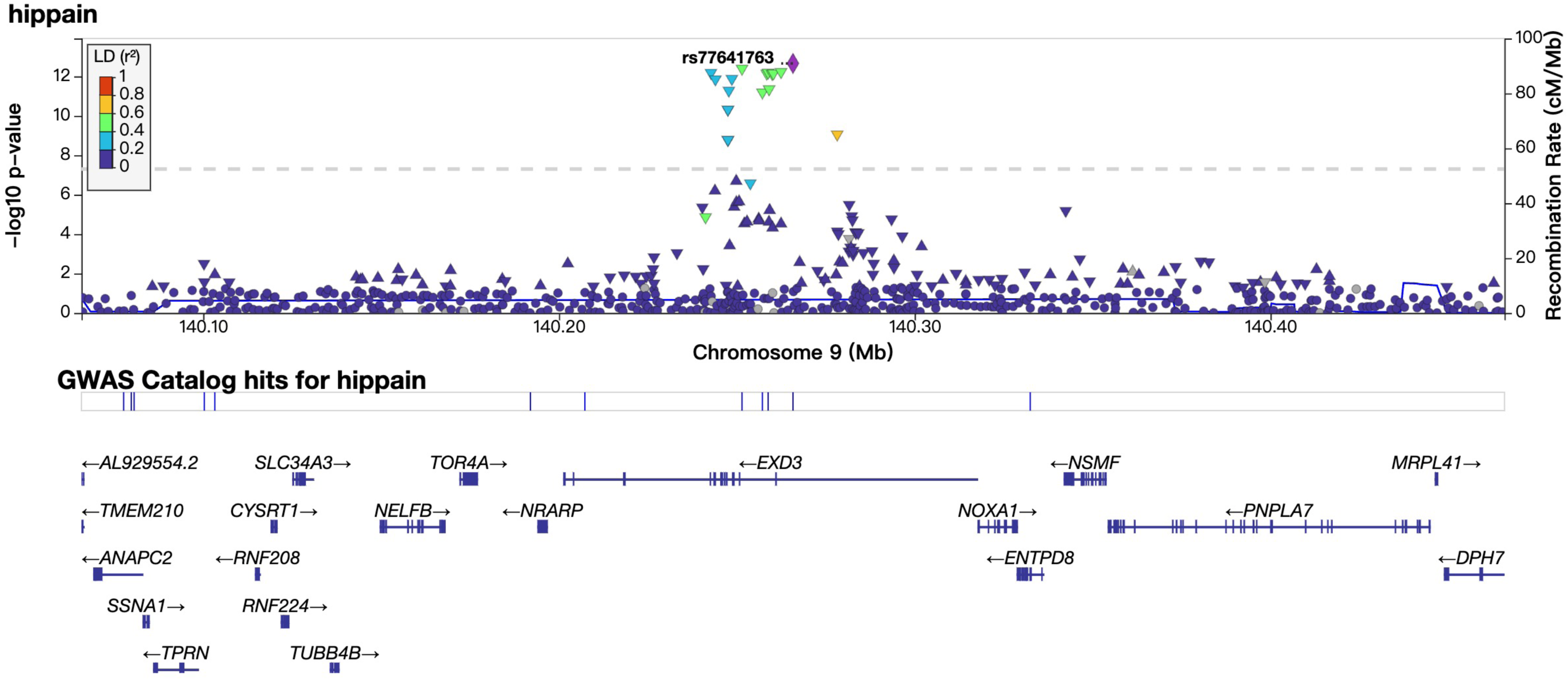
The regional plots of locus in *EXD3* region

**Table 2.** The 12 independent and significant SNPs within 7 loci identified by the GWAS on hip pain.

| Genomic Locus | rsID | Chr | SNP position | Effect allele | Discovery Stage (UK Biobank) |  |  |  | Replication Stage (Henkel et al. 2023) |  |
| --- | --- | --- | --- | --- | --- | --- | --- | --- | --- | --- |
|  |  |  |  |  | Non effective allele | Frequency the effect allele | Odds ratio | *p*-value | *p*-value in surgical hip OA | *p*-value in non-surgical hip OA |
| 1 | rs782595679 | 1 | 150340681 | GTT | G | 0.55 | 1.01 | 9.87x10<sup>-11</sup> | 0.37 | 0.057 |
| 1 | rs12042579 | 1 | 150454503 | C | T | 0.61 | 1.01 | 3.45x10<sup>-9</sup> | 2.30x10<sup>-4</sup> | 1.10x10<sup>-3</sup> |
| 2 | rs1422952 | 5 | 160806655 | A | T | 0.11 | 0.99 | 9.99x10<sup>-9</sup> | 0.14 | 0.78 |
| 2 | rs137863733 | 5 | 160890323 | CCACA | C | 0.79 | 1.01 | 1.30x10<sup>-8</sup> | 0.31 | 0.33 |
| 2 | rs62381584 | 5 | 160960864 | A | G | 0.98 | 1.03 | 3.51x10<sup>-8</sup> | 0.52 | 0.021 |
| 3 | rs77641763 | 9 | 140265782 | A | C | 0.87 | 0.99 | 2.20x10<sup>-13</sup> | 0.53 | 0.67 |
| 3 | 9:140247497_A_C | 9 | 140247497 | C | T | 0.88 | 0.99 | 4.79x10<sup>-11</sup> | 0.45 | 0.89 |
| 4 | rs761084711 | 10 | 61059104 | GC | G | 0.86 | 1.01 | 3.54x10<sup>-8</sup> | 0.65 | 0.67 |
| 5 | rs1330745 | 13 | 59159949 | C | T | 0.78 | 1.01 | 5.98x10<sup>-9</sup> | 0.23 | 0.1 |
| 5 | rs9597759 | 13 | 59122301 | A | C | 0.78 | 1.01 | 1.43x10<sup>-8</sup> | 9.80x10<sup>-3</sup> | 0.38 |
| 6 | rs2018384 | 16 | 71940825 | T | C | 0.26 | 1.01 | 4.77x10<sup>-8</sup> | 0.21 | 0.024 |
| 7 | rs12150561 | 17 | 42246196 | T | C | 0.95 | 1.02 | 4.09x10<sup>-9</sup> | 0.26 | 0.67 |

Chr: chromosome
OA: osteoarthritis
rs782595679, rs1422952, rs137863733, rs1330745 and rs2018384 were ascertained from cohorts contributing to the GWAS meta-analysis, specifically excluding data from the UK Biobank.

### Gene, gene-set and tissue expression analysis by FUMA

In our gene analysis, SNPs located within genes were mapped to 19,023 protein-coding genes. Consequently, the cut-off for genome-wide significance was established at *p* = 0.05/19,023 = 2.60 x 10^-6^. Notably, *EXD3* exhibited the most robust association, evidenced by a *p* value of 2.2 x 10^-13^. A total of seventeen genes demonstrated associations with hip pain, at *p* < 2.60 x 10^-6^ and these genes were *EXD3, RPRD2, TARS2, ECM1, PRPF3, C1orf51, MRPS21, C17orf53, ASB16, GABRB2, FAM13C, IST1, ATXN7L3, UBTF, ZNF821, ATXN1L* and *AP1G1*. Detailed results can be found in Supplementary Table 2.

In the gene-set analysis, we examined a total of 15,485 gene sets. Accordingly, the threshold for genome-wide statistical significance was set at *p* < 0.05 / 15,485 = 3.23 x 10^-6^. No gene sets were found to be statistically significant. The top ten gene sets from this analysis are presented in Supplementary Table 3.

In the tissue expression analysis, the overarching category of "Brain" was identified to have statistical significance across 30 general tissue types within certain organs. Notably, both "Brain_Cerebellum" and "Brain_Cerebellar_Hemisphere" demonstrated significant associations (*p* < 0.001) among 53 specific tissue types sourced from various organs. Further details and visualization of these results are available in Figure 3, 4.

**Fig. 3.**
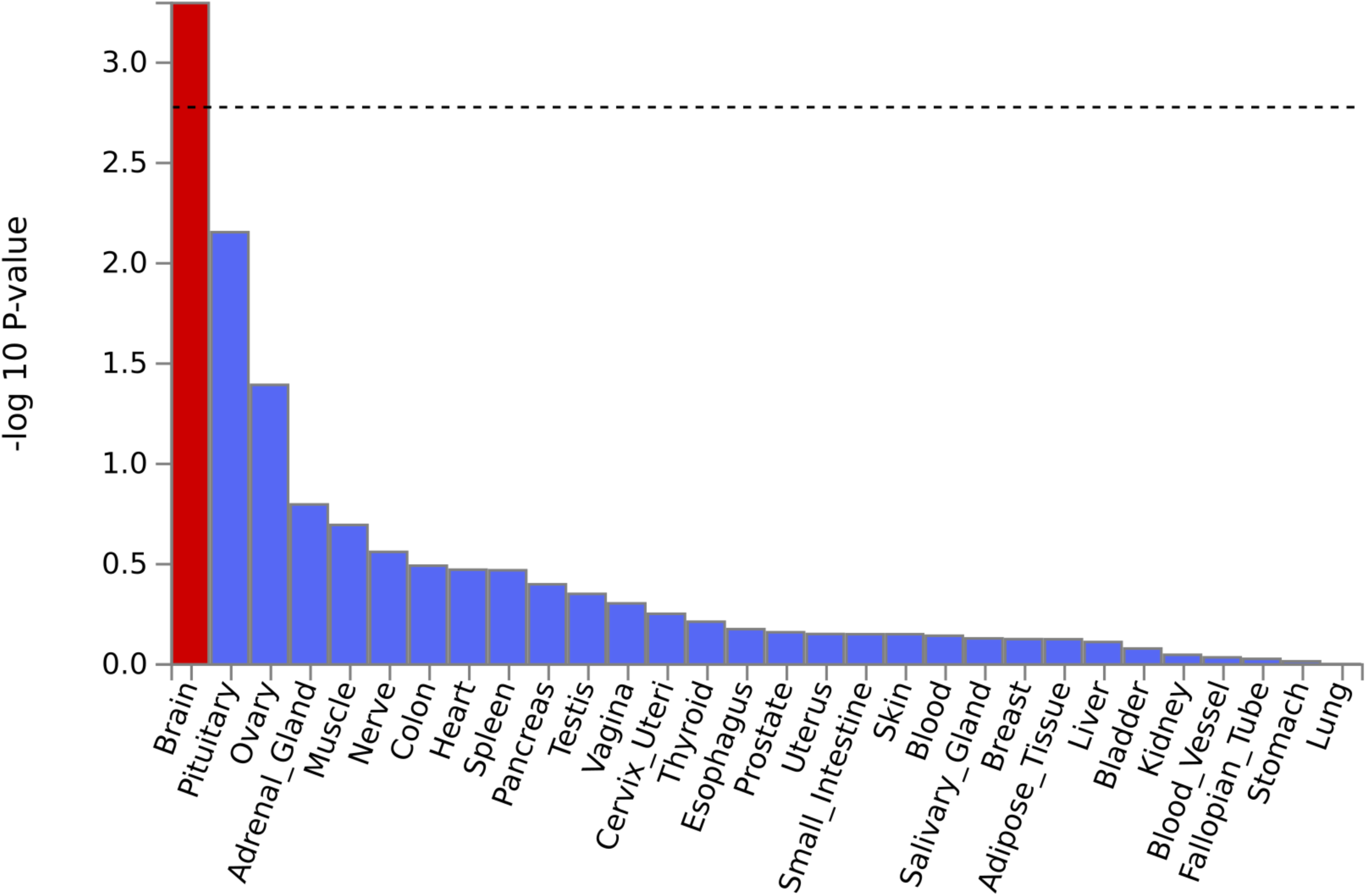
Tissue expression results on 30 specific tissue types by GTEx in the FUMA The dashed line shows the cut-off *p* value for significance with Bonferroni adjustment for multiple hypothesis testing

**Fig. 4.**
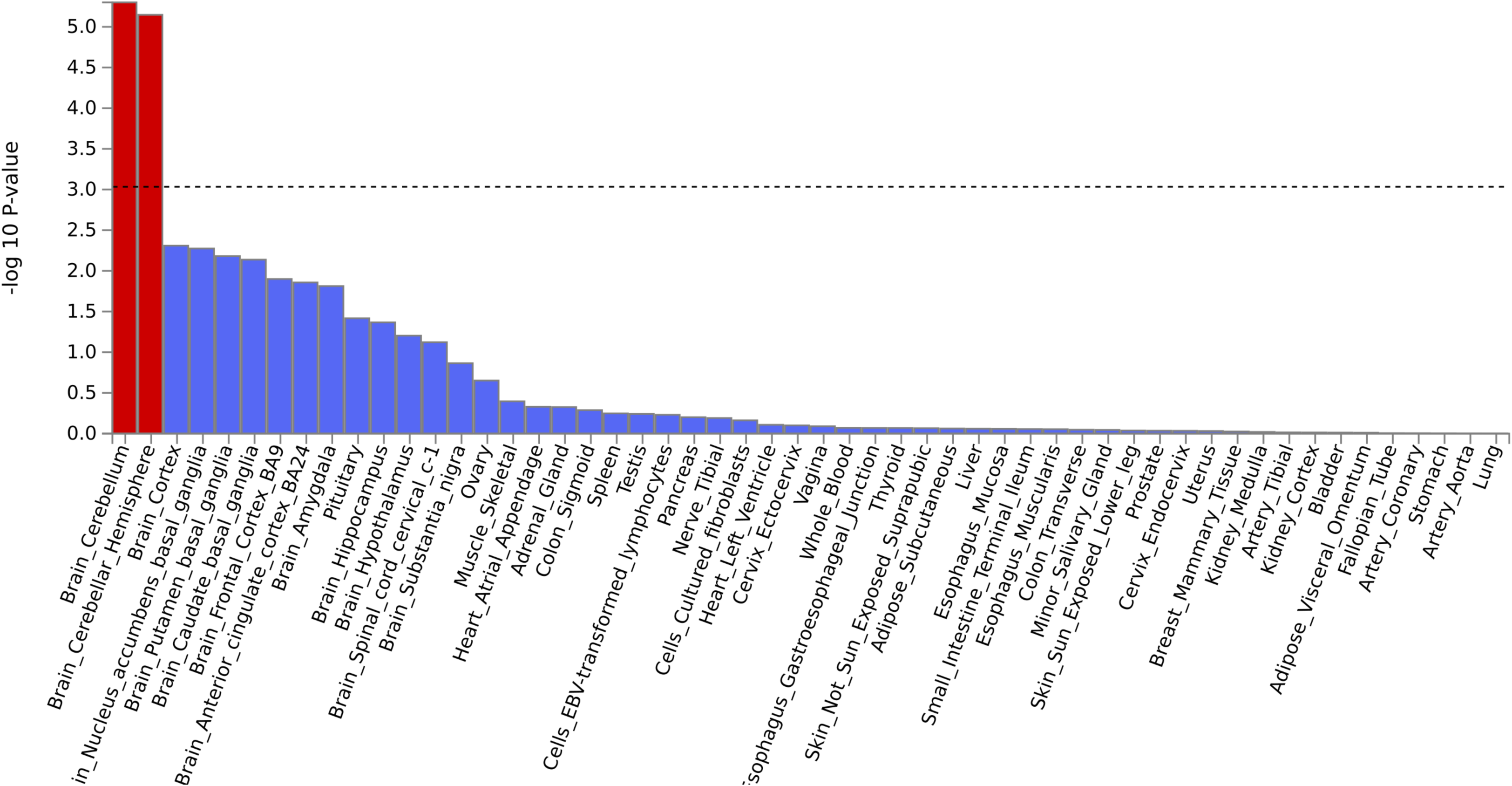
Tissue expression results on 53 specific tissue types by GTEx in the FUMA The dashed line shows the cut-off *p* value for significance with Bonferroni adjustment for multiple hypothesis testing

### Genetic correlation analysis by LDSC

In our exploration of the association between hip pain and other pain phenotypes, as well as the correlation between hip pain and hip OA (both surgical and non-surgical cases), we identified several noteworthy genetic correlations, as outlined in Table 3. After adjusting for multiple testing, the three most salient genetic correlations (*r_g_*) were as follows: Neck or shoulder pain (*r_g_* = 0.93, *p* = 3.96 x 10^-297^), multisite pain (*r_g_* = 0.92, *p* = 1 x 10^-14^) and back pain (*r_g_* = 0.80, *p* = 7.36 x 10^-^ 92).

**Table 3.** The significant genetic correlation results by LDSC between hip pain with other phenotypes.

| Trait 1 | Trait 2 | $r_g$ | $p$-value |
| --- | --- | --- | --- |
| Hip pain | Knee pain | 0.45 | $1.07 \times 10^{-13}$ |
| Hip pain | Multi-site pain | 0.92 | $1.00 \times 10^{-14}$ |
| Hip pain | Headache | 0.68 | $3.23 \times 10^{-201}$ |
| Hip pain | Facial pain | 0.60 | $1.36 \times 10^{-13}$ |
| Hip pain | Neck or shoulder pain | 0.93 | $3.96 \times 10^{-297}$ |
| Hip pain | Abdominal pain | 0.70 | $2.72 \times 10^{-12}$ |
| Hip pain | Back pain | 0.80 | $7.36 \times 10^{-92}$ |
| Hip pain | Non-surgical hip OA | 0.51 | $6.45 \times 10^{-24}$ |
| Hip pain | Surgical hip OA | 0.35 | $3.98 \times 10^{-23}$ |

$r_g$ : genetic correlation

## Discussion

In this GWAS of hip pain, which was conducted utilizing the comprehensive UK Biobank dataset, our team discerned noteworthy genetic variants across 7 distinct loci associated with self-reported hip pain. These findings highlight the complex genetic underpinnings associated with hip pain. Furthermore, our in-depth analysis illuminated clear genetic correlations between hip pain and several other pain phenotypes. Most notably, these correlations were observed with back pain and pain localized in the neck and shoulder regions. This suggests potential shared genetic pathways or mechanisms across these pain-related phenotypes. In this study, we employed the generic pain questionnaire from the UK Biobank, a valuable screening instrument, to ascertain the potential genetic underpinnings of heterogeneous pain phenotypes, such as hip pain. Using the UK Biobank for investigating heterogeneous phenotypes provides researchers with the advantage of mitigating potential issues related to reduced power due to heterogeneity. This is achieved by drawing on substantial sample sizes, thereby enhancing the clarity of statistical results amidst potential noise.

In this GWAS, we have analyzed and subsequently identified 7 distinct loci that were associated with hip pain. Among these, the most significant locus was situated within the *EXD3* gene on chromosome 9q34.3. This particular locus exhibited an impressively low *p* value of 2.20 x 10^-13^ for the marker rs77641763. This locus spans an extensive 37 kb within the *EXD3* gene and encompasses as many as 20 genome-wide significant SNPs, as detailed in Supplementary Table 1. The *EXD3* gene (Exonuclease 3’-5’ Domain Containing 3) is a member of the karyopherin family. This gene is pivotal in the intricate process of nuclear export, specifically facilitating the transport of certain cargo proteins from the nucleus to the cytoplasm (Liu et al. 2011). Such a mechanism is not merely a cellular routine but is of paramount importance in ensuring the correct cellular localization. This, in turn, is fundamental for a myriad of cellular functions, ranging from cell signaling to growth and differentiation (Finnegan and Pasquinelli 2013). While the protein encoded by the *EXD3* gene is recognized for its involvement in molecular transport through nuclear pore complexes in the nuclear envelope, a direct and unequivocal association between the *EXD3* gene and hip physiology remains elusive. However, delving deeper into cellular mechanisms, it becomes plausible to hypothesize that the indirect effects of genes, especially those involved in protein transport like *EXD3*, might have far-reaching impacts on various tissues. This includes the hip, where the modulation of the function of other genes or proteins integral to hip physiology could be influenced. Furthermore, it is worth noting that recent scientific investigations have unveiled an association between the *EXD3* gene and back pain (Freidin et al. 2019). Given the observed associations across different pain phenotypes, including both hip and back pain (Freidin et al. 2019), this cumulative evidence might not only bolster the hypothesis of the *EXD3* gene’s relevance to hip pain but also pave the way for future research in this domain.

The second SNP cluster was identified within the *RPRD2* gene located on chromosome 1, exhibiting a minimal *p* value of 9.87 x 10^-11^ for rs782595679. This locus spans 244 kb and encompasses 175 genome-wide significant SNPs, as detailed in Supplementary Table 1. The *RPRD2* gene, formally known as the Regulation of Nuclear Pre-mRNA Domain Containing 2, is implicated in the regulation of RNA polymerase II. This enzyme is pivotal in the transcription of DNA to synthesize precursor mRNA (Ni et al. 2014). Although the precise function of the protein encoded by the *RPRD2* gene remains a topic of ongoing research, its role in transcriptional regulation indicates its potential significance in various cellular processes (Ni et al. 2011). Considering the complex interplay of genes and proteins essential for the upkeep and repair of joint tissues, disruptions or alterations in the *RPRD2* gene might affect joint health. Within the realm of hip disorders, particularly hip OA, genes associated with transcriptional regulation could influence the equilibrium between cartilage synthesis and degradation. A 2019 GWAS study, also utilizing the UK Biobank data, established a correlation between the *RPRD2* gene and hip OA (Tachmazidou et al. 2019). Cumulatively, the *RPRD2* gene exhibits a discernible association with hip osteoarthritis supported by compelling biological evidence. Based on our research findings, it is postulated that the correlation is primarily attributed to the detection of hip pain emanating from osteoarthritis, rather than from other underlying pathologies.

Among the remaining GWAS loci in our analysis, we identified a significant locus on chromosome 5 situated in proximity to the *GABRB2* gene. Multiple SNPs were observed at this locus. The *GABRB2* gene encodes the beta-2 subunit of the gamma-aminobutyric acid (GABA) type A receptor, a principal inhibitory neurotransmitter receptor in the brain (Barki and Xue 2022). These receptors operate as ligand-gated chloride channels and are instrumental in inhibitory neurotransmission within the central nervous system (Yeung et al. 2018). Considering that chronic pain is frequently associated with neural signaling imbalances, the *GABRB2* gene could potentially influence its pathophysiology. Notably, several GWAS studies based on the UK Biobank have demonstrated a correlation between this gene and multisite chronic pain (Johnston et al. 2019; Johnston et al. 2021). Given the observed and calculated association between hip pain and other pain phenotypes, this locus warrants further investigation and research.

In addition, the genes *FAM13C, CTAGE16P, IST1*, and *ASB16* have emerged with considerable significance in our study. Notably, these genes had not been linked to hip disorders in earlier genetic investigations. Nevertheless, they are characterized by the presence of multiple significant SNPs, underscoring their potential importance. These specific regions are densely populated with genes. Furthermore, the aforementioned genes, *FAM13C*, *IST1*, and *ASB16*, are intricately involved in cellular processes and protein interactions (Bajorek et al. 2009; Burdelski et al. 2017; Fu et al. 2021; Liu et al. 2020). This involvement positions them as potential key players in the onset or progression of hip disorders. It is imperative, however, to emphasize that these preliminary observations necessitate rigorous further research to validate and elucidate their precise roles in hip disorders.

In our research, the hip pain phenotype we defined was a broad phenotype while the genetic variants we identified were postulated to be associated with hip osteoarthritis. Contemporary research has underscored the inherent association between hip pain and osteoarthritis, positing a possible genetic linkage between the two conditions. A comprehensive network meta-analysis underscored the efficacy of non-steroidal anti-inflammatory drugs and opioids in mitigating pain related to hip osteoarthritis, alluding to the intricate relationship between pain and this degenerative ailment (da Costa et al. 2021). The guidelines set forth by the American Physical Therapy Association further underscore the confluence of hip pain and mobility deficits, proposing that pain is not merely a symptom but a phenotype intrinsically connected to hip osteoarthritis (Cibulka et al. 2017). Notably, the hospital-diagnosed osteoarthritis cases (N = 10,083), self-reported osteoarthritis cases (N = 12,658) and hospital-diagnosed hip osteoarthritis (N = 2,397) from the UK Biobank were all much fewer in number than the number of the cases self-reporting hip pain in our study (N = 48563) (Zengini et al. 2018). This highlights the significance of addressing hip pain as an independent entity rather than merely as a symptom of hip disorders. In our assessment of the genetic correlation between hip pain and hip OA, the value for surgical hip OA was r_g_ = 0.35 and non-surgical hip OA was r_g_ = 0.51. In addition, 2 loci (*RPRD2*, *CTAGE16P*) were suggestively replicated in our study. This not only demonstrates the genetic similarity between hip pain and hip OA, but also suggests that hip pain can have its own loci as an independent phenotype. Furthermore, drawing upon our research findings, there exists a genetic correlation between hip pain and several other pain phenotypes, suggesting that shared genetic determinants underpinning these conditions. The top three pain phenotypes most genetically correlated with hip pain were neck or shoulder pain (*r_g_* = 0.93), multisite pain (*r_g_* = 0.92) and back pain (*r_g_* = 0.80). Our study demonstrates an interplay of genetic determinants in the manifestation of pain, corroborating previous research. Specifically, a prior network analysis has identified a substantial cluster of conditions and pinpointed arthropathic, back, and neck pain as potential hubs for cross-condition chronic pain (Zorina-Lichtenwalter et al. 2023).

Gene analysis conducted through FUMA also supported our findings, highlighting *EXD3, RPRD2*, and *GABRB2* as significant genes associated with hip pain. Furthermore, the tissue expression analysis facilitated by FUMA indicated a connection between the “Brain_Cerebellum” pathway and the phenotype under study. Notably, our research determined the SNP-based heritability for hip pain to be 0.12. Upon comparing this outcome with the previously reported heritability figure of 0.08 attributed to knee pain, a similarity in heritability manifests between the two distinct pain phenotypes is seen. (Meng et al. 2019).

Our research, though providing initial insights, encounters specific limitations in consolidating the validation phase of our results. In the replication phase, we utilized a different phenotype (hip OA) instead of hip pain, given that hip pain is a prominent indicator of hip OA. This deviation was necessitated by the absence of datasets on hip pain for the replication phase. Additionally, although we highlighted replication results derived from cohorts exclusive of the UK Biobank, it is important to acknowledge that the statistical power of these results may be comparatively limited. This is primarily because the sample size of UK Biobank cohort is larger than the other two cohorts included in the replication dataset. An additional limitation pertains to the selection validity of the control group in our research. While we used data from participants who reported no pain as the control cohort, this approach may inadvertently identify loci that are associated with pain generally, rather than correlated with hip pain. This issue has been partially mitigated through the utilization of genetic correlation analysis between pain sites, which shows that there are both similarities and differences in the genetic architecture of hip pain and other pains. For readers’ interest, we have included the results of the GWAS contrasting hip pain versus no hip pain (rather than no pain) in Supplementary Figure 4, which features the Manhattan plot. Furthermore, it is essential to note the methodological approach adopted in our study. We characterized hip pain cases and controls by relying on the responses provided by the UK Biobank participants to a specific question. This question was designed to gauge the occurrence of hip pain significant enough to disrupt or interfere with daily activities within the past month. However, the question did not delve into further details, such as the severity of the pain, or the frequency with which it occurred, or the exact anatomical location of the pain within the hip region. Given this, while the question provides some insights, it does so only at a high level. Consequently, the phenotype definition that we derived based on these responses should be perceived and interpreted as being broadly defined. Future research may need to incorporate more detailed questionnaires and larger sample sizes to provide a comprehensive understanding of the topic.

## Conclusion

In summary, our study suggested 7 new genetic loci which are associated with hip pain, shedding further light on its potential mechanisms. Through GWAS, we have identified these loci that have not been previously linked to hip pain, expanding the current understanding of the genetic underpinnings of this condition. Future studies are warranted to validate these associations in diverse populations and to explore the potential interactions between these genetic loci and other factors.

## Data Availability

All data produced in the present study are available upon reasonable request to the authors
All data produced in the present work are contained in the manuscript
All data produced are available upon request

## Statements and Declarations

### Funding

This study was mainly funded by the Pioneer and Leading Goose R&D Program of Zhejiang Province 2023 with reference number 2023C04049 and Ningbo International Collaboration Program 2023 with reference number 2023H025. This work was also supported by the European Union’s Horizon 2020 research and innovation programme under grant agreement No 633491 (DOLORisk).

### Data availability

This study will adhere to all ethical guidelines and data protection protocols of the UK Biobank. The current study was conducted under approved UK Biobank data application number 89386. The summary statistics of the UK Biobank results on hip pain are available upon request. Any other data relevant to the study that are not included in the article or its supplementary materials are available from the authors upon reasonable request.

### Authors’ Contributions

QP drafted the paper and performed the UK Biobank GWAS analysis. TC contributed to data formatting. AV, HLH, PZ, MH, TD, LAC and BLH provided comments to the paper. WM organised the project and provided comments.

### Corresponding authors

Correspondence to Weihua Meng

### Consent to Publish

All authors provide consent for publication.

### Ethical Approval

This study was approved by the Ethics Committee of the University of Nottingham Ningbo China.

## Notes

### Competing Interest Statement

The authors have declared no competing interest.

### Funding Statement

This study was funded by the Pioneer and Leading Goose R&D Program of Zhejiang Province 2023 with reference number 2023C04049 and Ningbo International Collaboration Program 2023 with reference number 2023H025.
This work was also supported by the European Union's Horizon 2020 research and innovation programme under grant agreement No 633491 (DOLORisk).

### Author Declarations

The dataset we used was approved by the UK Biobank with a project number 89386. The ethics of this UK Biobank project was approved by the ethical committee of the University of Nottingham Ningbo China.

