## Supplementary figures and images for "A genome-wide association study identifies genetic variants associated with hip pain in the UK Biobank cohort (N=221,127)"

### Supplementary Figure 1

#### Slide 1
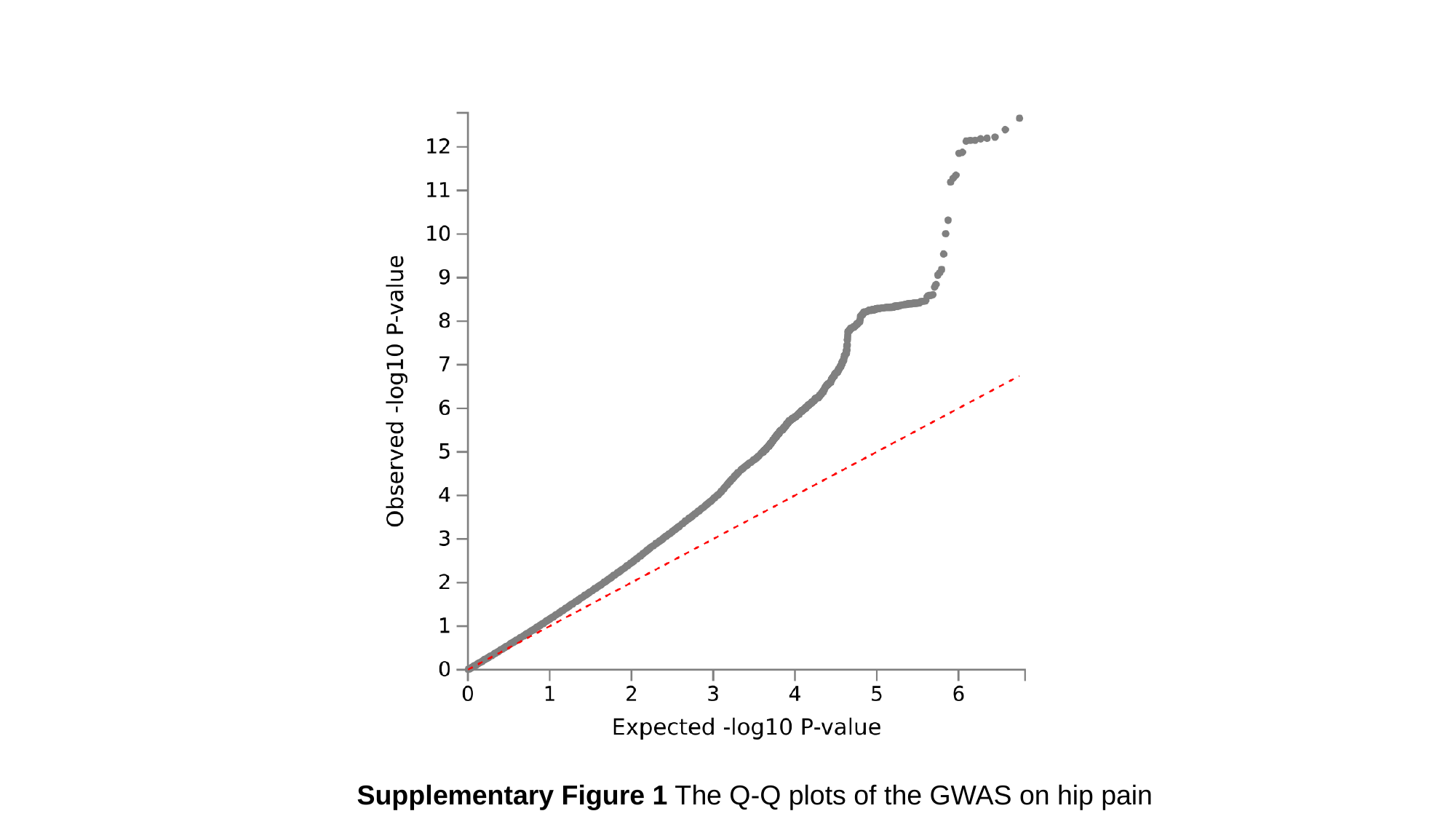

Supplementary Figure 1 The Q-Q plots of the GWAS on hip pain

### Supplementary Figure 2

#### Slide 1
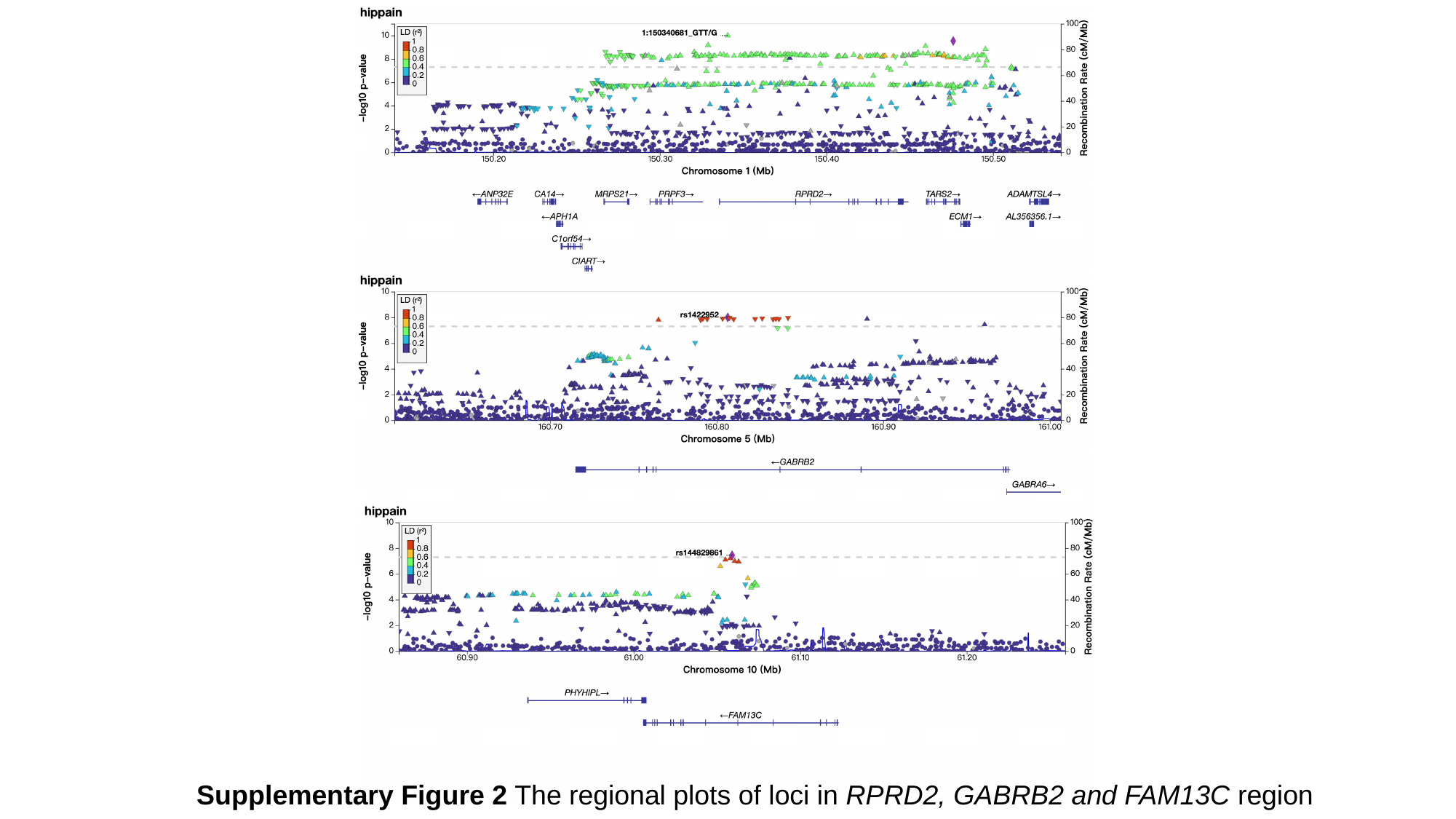

Supplementary Figure 2 The regional plots of loci in RPRD2, GABRB2 and FAM13C region

### Supplementary Figure 3

#### Slide 1
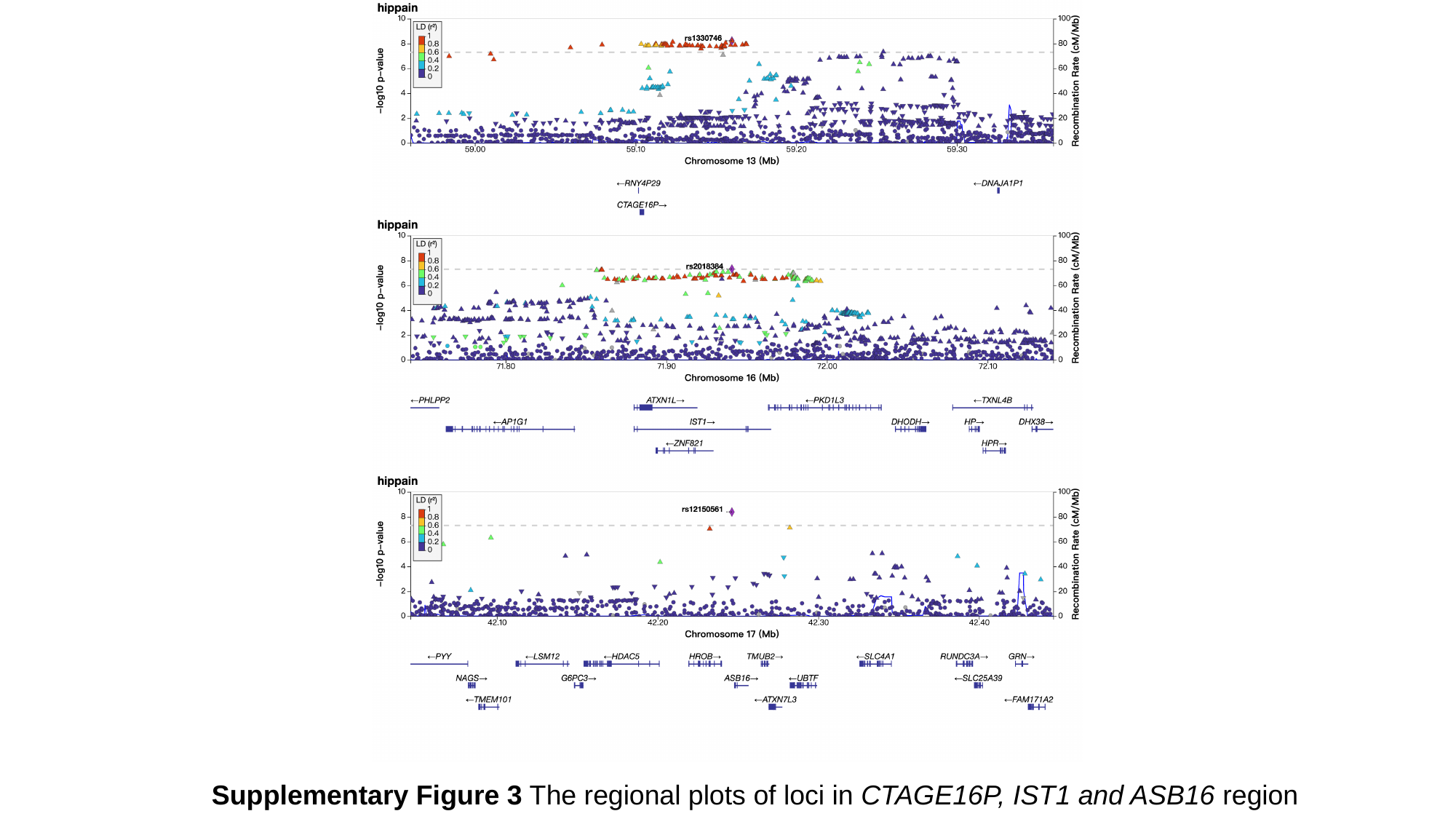

Supplementary Figure 3 The regional plots of loci in CTAGE16P, IST1 and ASB16 region
