## Supplementary Figure 4 for "A genome-wide association study identifies genetic variants associated with hip pain in the UK Biobank cohort (N=221,127)"

### Slide 1
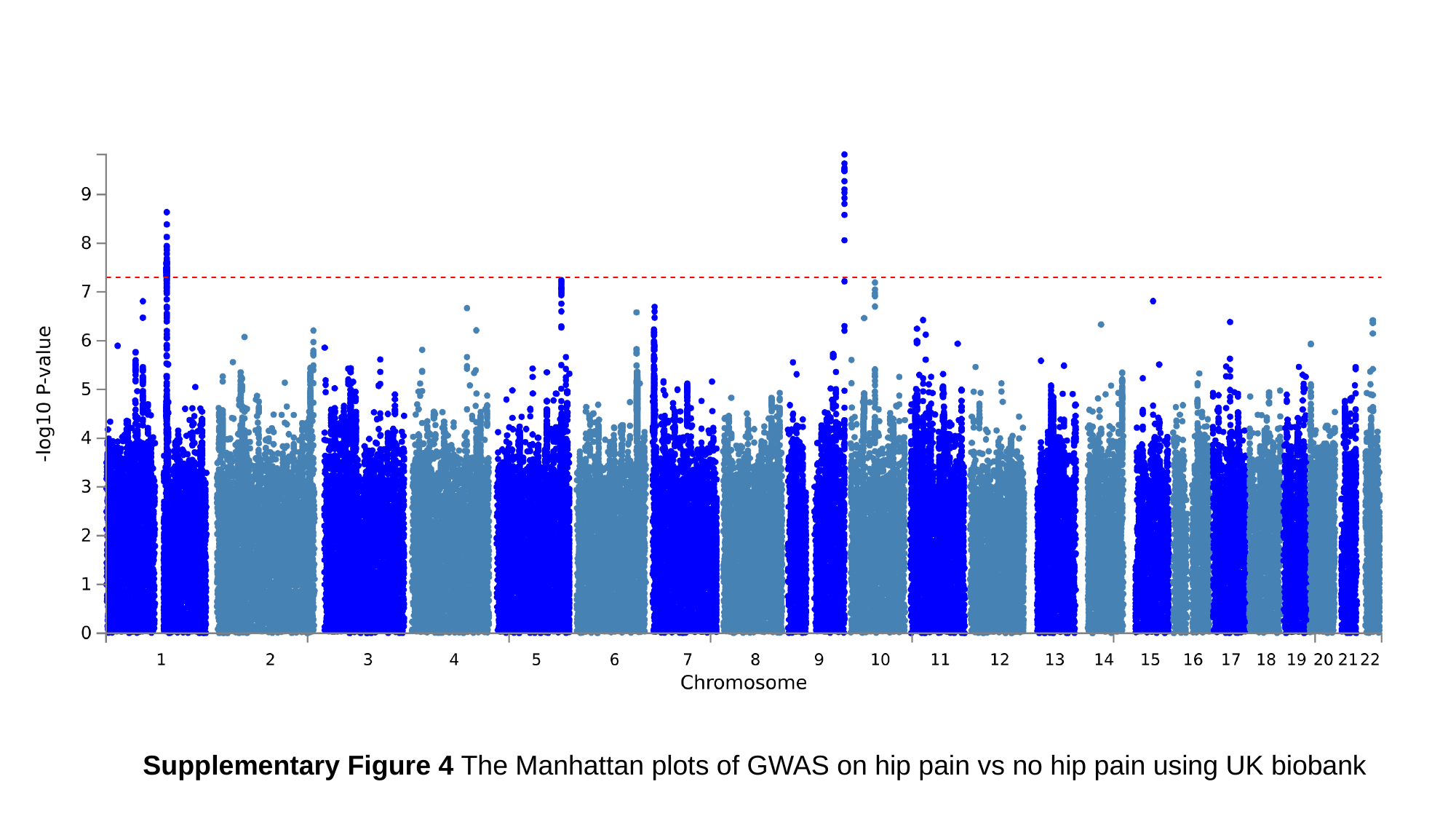

Supplementary Figure 4 The Manhattan plots of GWAS on hip pain vs no hip pain using UK biobank
